# Did social factors buffer against the effect of adversities on self-harm during the COVID-19 pandemic? A longitudinal analysis of 49,227 UK adults

**DOI:** 10.1101/2021.06.19.21259173

**Authors:** Elise Paul, Daisy Fancourt

## Abstract

**Background:** Little is known about which factors exacerbate and buffer the impact of COVID-19 -related adversities on changes in thinking about and engaging in self-harm over time.

**Aims:** To examine how changes in four social factors contribute to changes in self-harm thoughts and behaviours over time and how these factors in turn interact with adversities and worries about adversities to increase risk for these outcomes.

**Method:** Data from 49,227 UK adults in the UCL COVID-19 Social Study were analysed across the first 59 weeks of the pandemic. Fixed effects logistic regressions examined time-varying associations between social support quality, loneliness, number of days of face-to-face contact for ≥15 minutes, and number of days phoning/video calling for ≥15 minutes with self-harm thoughts and behaviours. We then examined how these four factors in turn interacted with the total number of adversities and worries about adversity on outcomes.

**Results:** Increases in the quality of social support decreased the likelihood of both outcomes, whilst greater loneliness increased their likelihood. Associations were inconsistent for telephone/video contact and face-to-face contact with outcomes. Social support buffered and loneliness exacerbated the impact of adversity experiences with self-harm behaviours. Other interactions were inconsistent, and some were in the unexpected direction.

**Conclusions:** These findings suggest the importance of the quality of one’s social support network, rather than the mere presence of contact, is important for reducing the likelihood of self-harm behaviours in the context of COVID-19 pandemic-related adversity and worry.

## Introduction

There is concern that the COVID-19 pandemic as well as its economic aftermath will have a negative impact on suicides, although this is not inevitable.^1,2^ This is hypothesised to occur via reductions in protective factors such as social connectedness and increases in risk factors such as domestic abuse and unemployment.^2–4^ According to the diathesis-stress models of suicide risk, the experience of these near-term stressors for individuals who already have enduring risk factors (such as traits of impulsivity, genetic vulnerability, and having experienced adversity early in life) could lead to an increase in self-harm behaviours and ultimately suicide.^5–7^

There is already evidence that the pandemic is having a detrimental effect on self-harm thoughts and behaviours, which significantly increase the probability of eventual death by suicide.^8,9^ Although early in the pandemic there was a reduction in the number of clinical presentations for self-harm compared to prior years, this could have been due to a decrease in face-to-face services and a wish to protect health care services.^10^ However, survey data suggest that a greater proportion of the population are thinking about or actually harming themselves than pre-pandemic.^11–13^ Stressful life events which can precipitate self-harm thoughts and behaviours in the short term such as domestic abuse^14^ and widespread unemployment^15,16^ have also increased. Concerns and worries about these and other adverse events that can be proximal triggers for self-harm^6,7^ have been found to have a similar impact on self-harm thoughts and behaviours as actually experiencing these adversities during the COVID-19 pandemic.^17^ Further, the combination of the unprecedented social distancing requirements, uncertainty about the future, and the accumulation of stressful circumstances have the potential to increase risk for suicides as the pandemic wears on and in the expected upcoming economic recession as well.^2,3^

There are, however, factors that could protect against the impact of these stressful circumstances on self-harm thoughts and behaviours. More frequent contact with others as well as the quality of one’s social support has been shown to provide a buffer against the likelihood of self-harm in the context of acute stressors such as financial difficulties or relationship breakdown.^18,19^ Given that the most commonly cited reasons for self-harm are to relieve suffering and manage distress,^20,21^ it follows that having access to supportive, understanding others would help mitigate the adverse consequences occurring in the context of the pandemic. However, social restrictions imposed in the current circumstances may have severely limited access to drawing on and maintaining connections vital to reducing the impact of this stress. Therefore, an unresolved question is whether the perceived quality of one’s social support and more frequent contact with others buffer the impact of adversity and worry about adversity on thinking about and engaging in self-harm.

A second factor that may be important for the link between pandemic-related adversities and self-harm thoughts and behaviours is loneliness,^2^ or the subjective distress resulting from a discrepancy between desired and perceived social relationships.^22^ Loneliness and a lack of social integration have been emphasised as important factors for suicide from Durkheim’s early sociological studies of suicide^23^ to modern theories of suicide risk.^6,7^ Whilst early in the pandemic, loneliness did not seem to be higher than pre-pandemic levels,^18,24^ recent data from the Office of National Statistics’ Opinions and Lifestyle Survey in the UK indicate that over the past year, the proportion of people who are lonely ‘often’ or ‘always’ has increased (5.0% to 7.2%).^26^ Thus, risk for self-harm thoughts and behaviours due to adversities and worries about adversity may be exacerbated in the present circumstances by increased levels of loneliness.

In sum, although the COVID-19 pandemic is having a detrimental impact on a number of known risk factors for suicide, little is known about how social factors such as social support, social contact, and loneliness may interact with these risk factors to exacerbate or buffer their impact on self-harm thoughts and behaviours. The aim of this study is therefore to establish which near-term social factors are associated with changes over time in self-harm thoughts and behaviours in a large sample of UK adults across the first 59 weeks of the COVID-19 pandemic. Specifically, we explore the time-varying longitudinal relationships between i) the quality of one’s social support, ii) loneliness, iii) time spent in face-to-face contact with others, and iv) time spent in phone or video contact with others with changes in these two outcomes. We then also examine whether these factors moderate the relationship between experiencing and worrying about adversities and self-harm thoughts and behaviours. We improve upon prior research in this area by using fixed effects statistical modelling methodology^27^ which accounts for longer-term more stable risk factors for self-harm thoughts and behaviours such as genetic predisposition and certain personality traits.^5,7^ The findings will further our understanding of which factors attenuate and exacerbate the risk for self-harm thoughts and behaviours in the context of the COVID-19 pandemic which are important for informing suicide prevention efforts.

## Methods

### Participants

We used data from the COVID-19 Social Study; a large ongoing panel study of the psychological and social experiences of over 70,000 adults (aged 18+) in the UK during the COVID-19 pandemic. The study commenced on 21 March 2020 and involves online weekly (from August 2020, monthly) data collection across the pandemic. Sampling is not random and therefore is not representative of the UK population, but the study does contain a heterogeneous sample. The sample was recruited using three primary approaches. First, convenience sampling was used, including promoting the study through existing networks and mailing lists (including large databases of adults who had previously consented to be involved in health research across the UK), print and digital media coverage, and social media. Second, more targeted recruitment was undertaken focusing on (i) individuals from a low-income background, (ii) individuals with no or few educational qualifications, and (iii) individuals who were unemployed. Third, the study was promoted via partnerships with third sector organisations to vulnerable groups, including adults with pre-existing mental health conditions, older adults, carers, and people experiencing domestic violence or abuse.

The authors assert that all procedures contributing to this work comply with the ethical standards of the relevant national and institutional committees on human experimentation and with the Helsinki Declaration of 1975, as revised in 2008. The study was approved by the UCL Research Ethics Committee [12467/005] and all participants gave informed consent. The study protocol and user guide (which includes full details on recruitment, retention, data cleaning, and sample demographics) are available at https://github.com/UCL-BSH/CSSUserGuide.

For these analyses, we used data from the fourteen months between 01 April 2020 to 17 May 2021 (n = 66,308, observations = 918,440). Participants were eligible for inclusion in the analysis if they had three or more data collections over the study period (n = 52,569 [79.3%], observations = 899,447 [97.9%]). We excluded participants with missing data on any of the variables in the study. The final sample size is 49,227 (observations = 849,800). See Supplemental Table S1 for a comparison of excluded and included participants.

### Measures

#### Self-harm thoughts and behaviours

Thoughts of death or self-harm (hereafter *self-harm thoughts*) was measured with an item from the Patient Health Questionnaire (PHQ-9);^28^ an instrument often used as a diagnostic tool for depression in primary care practice: “Over the last week, how often have you been bothered by thoughts that you would be better off dead or hurting yourself in some way?”. Second, self-harm behaviours were measured with a similar study-developed item: “Over the last week, how often have you been bothered by self-harming or deliberately hurting yourself?”. Responses to both items were rated on a four-point scale from “not at all” to “nearly every day” and collapsed into binary variables indicating the presence of at least some self-harm thoughts or self-harm behaviours at each time point.

### Social Support

#### Perceived social support

Social support in the past week was measured using an adapted version of the six-item short form of Perceived Social Support Questionnaire (F-SozU K-6).^29,30^ Each item is rated on a 5-point scale from “not true at all” to “very true”, with higher scores indicating higher levels of perceived total social support (hereafter, ‘social support’). Minor adaptations made to question phrasing to make it relevant to experiences during COVID-19 can be found in Supplemental Table S2. We also disaggregated the total social support variable into emotional support and instrumental support (3 items each) to examine whether the provision of instrumental assistance or emotional support may have been driving any findings. Mean social support scores were calculated at each time point.

#### Time spent in contact with others

Two continuous variables representing the number of days participants i) had face-to-face contact with another person for 15 minutes or more (including someone the participant lives with), and ii) had a phone or video call with another person for 15 minutes or more in the past week were included. The mean for each of these variables was calculated at each time point.

### Loneliness

Loneliness was measured using the 3-item UCLA-3 Loneliness, a short form of the Revised UCLA Loneliness Scale (UCLA-R).^31^ Each item is rated with a 3-point rating scale, ranging from “hardly ever” to “often”. The mean of these three items was calculated for each participant and higher scores indicate higher loneliness.

### Adversity experiences and worries

#### Adversity experiences

Five categories of adversities occurring in the past week were considered: financial adversity, COVID-19 illness, family/friend serious illness or bereavement, experiencing physical or psychological abuse, and not being able to access essential items. Each category of adversity was treated as binary (absent vs. present) and summed to create an index of the number of adversities experienced at each time point. More detailed description of these measures can be found in Table S3).

#### Worries about adversity

Five worries about adverse experiences were measured at the same time as the adversity measures and selected to correspond with these variables. Each category of worry was operationalised as binary (absent vs present): financial worries, worries about COVID-19 illness, social and relationship worries, concerns about safety and security, and worries about accessing essentials. These binary variables were then summed to create the total number of worries about adversity at each time point.

### Statistical analysis

We used separate fixed-effects regression models for each social variable to analyse the time-varying associations between changes in social support, time spent in contact with others face-to-face and via telephone/video, and loneliness with changes in self-harm thoughts and behaviours over the course of the study period (1 April 2020 to 17 May 2021). In the fixed effects approach, individuals serve as their own reference point which accounts for any confounding associations between time-invariant (stable) covariates such as socio-economic status, genetics, personality, and history of mental illness between predictors and outcomes.^32^ We then repeated these models including the total number of adversities and the total number of worries about adversity and the interactions between these variables and each of the four social variables in turn. See Supplementary Materials for more detail, including model equation. Models controlled for day of the week and number of days since the first UK lockdown commenced. Resulting regression coefficients were exponentiated and presented as odds ratios along with 95% confidence intervals.

To account for the non-random nature of the sample and increase representativeness of the UK general population, all data were weighted to the proportions of gender, age, ethnicity, country, and education obtained from the Office for National Statistics.^33^ Weights were constructed using a multivariate reweighting method using the Stata user written command ‘ebalance’.^34^ Analyses were conducted using Stata version 16.^35^

Sensitivity analyses with the social support variable disaggregated into emotional support and instrumental support (3 items each) were conducted to examine whether the provision of instrumental assistance or emotional support was driving any social support findings.

## Results

### Descriptive statistics

Descriptive statistics for the total sample and for those with any change in self-harm thoughts or self-harm behaviours) are presented in Tables S1 and Table S4, respectively. Before weighting, the total sample was disproportionately female, of older age, and highly educated (Table S1). After weighting, sample proportions reflected those of the UK population (Table S4). The average number of measurement points in the total sample was 22.66 (SD = 7.36). There was within-individual variation in both of the self-harm outcome measures, predictors, and adversity measures (Table 1). Nearly one-quarter (23.5%) of the sample reported self-harm thoughts at least once over the study period, while 7.6% reported self-harm behaviours at least once.

**Table 1:**
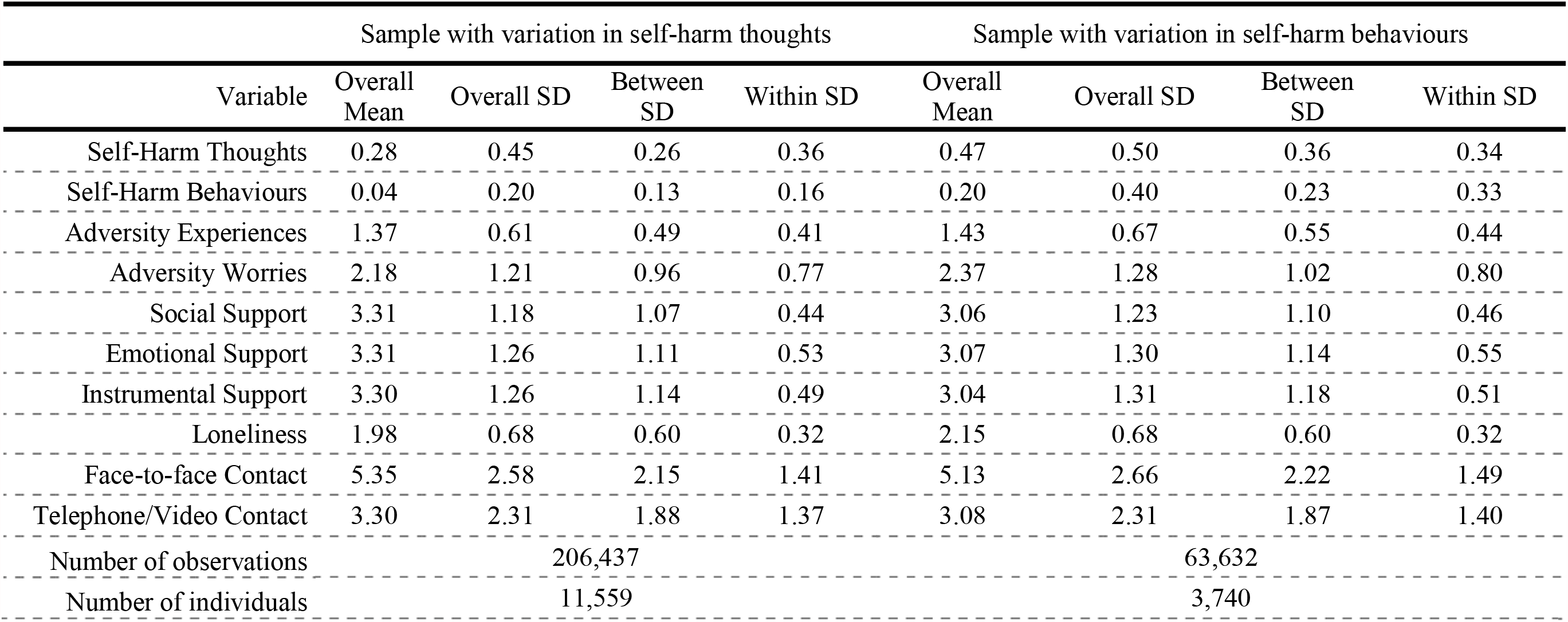
Means and standard deviations for study outcomes and exposures amongst individuals with variation in each outcome variable

### Associations between predictor variables and self-harm thoughts and behaviours

Better quality social support was associated with the largest reductions in the odds of self-harm thoughts (OR = 0.55; 95% CI = 0.54 to 0.57) and self-harm behaviours (OR = 0.71; 95% CI = 0.68 to 0.74), whilst loneliness associated with a 3.77 (95% CI = 3.61 to 3.93) times higher odds of self-harm thoughts and 2.18 (95% CI = 2.02 to 2.34) higher odds of self-harm behaviours (Table 2). The number of days in which individuals had had face-to-face contact or telephone/video contact with another person for at least 15 minutes were associated with small reductions in self-harm thoughts, but not self-harm behaviours.

**Table 2:**
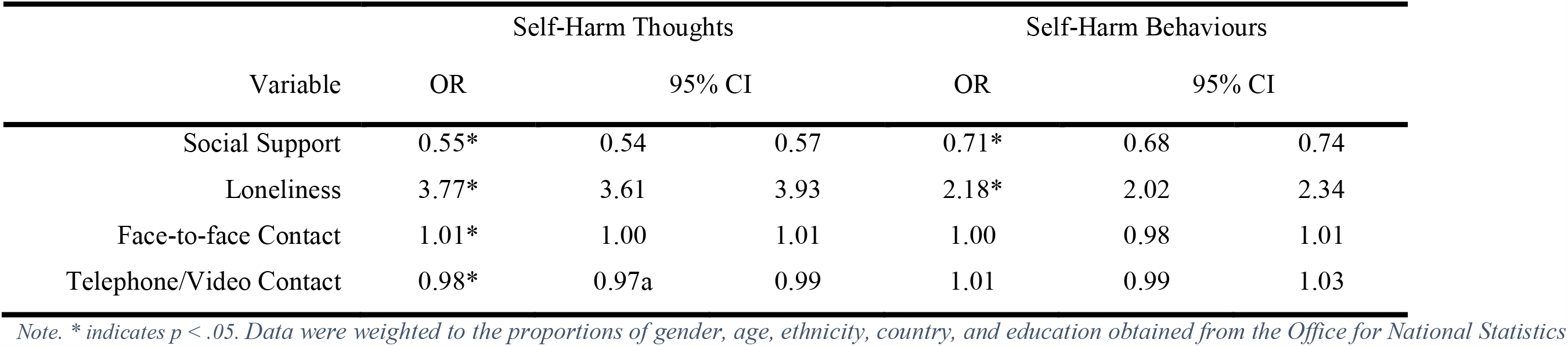
Associations between predictor variables with self-harm thoughts and behaviours (main effects) derived from fixed effects logistic regression models.

| Variable | Self-Harm Thoughts |  |  | Self-Harm Behaviours |  |  |
| --- | --- | --- | --- | --- | --- | --- |
|  | OR | 95% CI |  | OR | 95% CI |  |
| Social Support | 0.55* | 0.54 | 0.57 | 0.71* | 0.68 | 0.74 |
| Loneliness | 3.77* | 3.61 | 3.93 | 2.18* | 2.02 | 2.34 |
| Face-to-face Contact | 1.01* | 1.00 | 1.01 | 1.00 | 0.98 | 1.01 |
| Telephone/Video Contact | 0.98* | 0.97a | 0.99 | 1.01 | 0.99 | 1.03 |

Note. \* indicates $p < .05$ . Data were weighted to the proportions of gender, age, ethnicity, country, and education obtained from the Office for National Statistics

### Interactions between predictors and adversities and adversity worries

The main effects of predictor variables, adversities, and adversity worries, as well as their interactions are shown in Table 3. Main associations between adversity worries and both outcomes were generally larger in magnitude (OR range = 1.24 to 1.42) than for actual adversity experiences (1.08 to 1.14) with both outcomes. Social support and loneliness continued to associate with reduced and increased likelihood, respectively, of self-harm thoughts and behaviours, even when adversities and worries were included in the models.

**Table 3:** Associations between adversity experiences with self-harm thoughts and behaviours (main effects and interaction terms with predictor variables) derived from fixed effects logistic regression models.

| ADVERSITY EXPERIENCES | Self-Harm Thoughts |  |  | Self-Harm Behaviours |  |  |
| --- | --- | --- | --- | --- | --- | --- |
|  | OR | 95% CI |  | OR | 95% CI |  |
| Adversity Experiences | 1.13* | 1.11 | 1.15 | 1.10* | 1.06 | 1.13 |
| Social Support | 0.42* | 0.40 | 0.43 | 0.63* | 0.60 | 0.66 |
| Interaction w/Social Support | 1.00 | 0.99 | 1.02 | 0.97* | 0.95 | 0.99 |
| Adversity Experiences | 1.13* | 1.11 | 1.15 | 1.08* | 1.04 | 1.12 |
| Loneliness | 2.62* | 2.55 | 2.69 | 1.72* | 1.64 | 1.80 |
| Interaction w/Loneliness | 0.99 | 0.97 | 1.00 | 1.04* | 1.01 | 1.06 |
| Adversity Experiences | 1.13* | 1.12 | 1.15 | 1.13* | 1.10 | 1.16 |
| Face-to-face Contact | 0.94* | 0.92 | 0.96 | 0.96* | 0.92 | 0.99 |
| Interaction w/Face-to-face Contact | 0.99 | 0.98 | 1.00 | 1.00 | 0.99 | 1.02 |
| Adversity Experiences | 1.14* | 1.12 | 1.15 | 1.13* | 1.10 | 1.16 |
| Telephone/Video Contact | 0.91* | 0.89 | 0.93 | 0.99 | 0.95 | 1.02 |
| Interaction w/Telephone/Video Contact | 0.98 | 0.97 | 1.00 | 1.01 | 0.99 | 1.03 |
| **ADVERSITY WORRIES** |  |  |  |  |  |  |
| Adversity Worries | 1.40* | 1.37 | 1.43 | 1.31* | 1.25 | 1.36 |
| Social Support | 0.42* | 0.41 | 0.44 | 0.62* | 0.59 | 0.66 |
| Interaction w/Social Support | 1.01 | 0.99 | 1.02 | 1.03* | 1.00 | 1.06 |
| Adversity Worries | 1.40* | 1.36 | 1.43 | 1.24* | 1.19 | 1.31 |
| Loneliness | 2.60* | 2.52 | 2.67 | 1.69* | 1.61 | 1.78 |
| Interaction w/Loneliness | 0.95* | 0.94 | 0.97 | 1.00 | 0.97 | 1.03 |
| Adversity Worries | 1.42* | 1.39 | 1.45 | 1.30* | 1.26 | 1.35 |
| Face-to-face Contact | 0.93* | 0.91 | 0.95 | 0.95* | 0.92 | 0.99 |
| Interaction w/Face-to-face Contact | 1.02* | 1.00 | 1.03 | 1.02 | 0.99 | 1.04 |
| Adversity Worries | 1.42* | 1.39 | 1.45 | 1.31* | 1.27 | 1.36 |
| Telephone/Video Contact | 0.90* | 0.88 | 0.92 | 0.96 | 0.92 | 1.00 |
| Interaction w/Telephone/Video Contact | 0.98* | 0.96 | 0.99 | 1.04* | 1.02 | 1.07 |

There was evidence that the relationship between adversity worries and self-harm thoughts was slightly attenuated by lower levels of loneliness (OR = 0.95; 95% CI = 0.94 to 0.97) and increases in days of telephone/video contact (OR = 0.98; 95% CI = 0.96 to 0.99), but this relationship was slightly exacerbated by days of face-to-face contact (OR = 1.02; 95% CI = 1.00 to 1.03).

For self-harm behaviours, loneliness exacerbated (OR = 1.04; 95% CI = 1.01 to 1.06) the association of adversity experiences with this outcome, whilst better quality social support attenuated this association (OR = 0.97; 95% CI = 0.95 to 0.99). Social support and days of telephone/video contact exacerbated the relationship between adversity worries and self-harm behaviours.

### Sensitivity analyses

Sensitivity analyses with the disaggregated social support scale into emotional and instrumental support indicated slightly stronger associations between emotional support and self-harm thoughts (OR = 0.53; 95% CI = 0.51 to 0.55) than between instrumental support and self-harm thoughts (0.76; 95% CI = 0.74 to 0.79) (Table S5). But the differences between these two types of support were negligible in relation to self-harm behaviours. Other substantive findings for the main associations of adversities and adversity worries were the same as in the main analyses (Table S6). There was weak evidence that emotional support exacerbated the relationship between adversity worries and both outcomes. Instrumental support was the only variable to attenuate any of the associations (adversity experiences with self-harm behaviours).

## Discussion

Better quality social support considerably reduced the likelihood of both of self-harm thoughts and self-harm behaviours across the first 59 weeks of the COVID-19 pandemic. Having had more days of telephone/video contact with another person for 15 minutes or more led to only minor reductions in the likelihood of self-harm thoughts and was not associated with behaviours. Additionally, increases in loneliness were associated with a nearly 4-fold and over 2-fold likelihood of self-harm thoughts and behaviours, respectively. These findings suggest that the quality of social interactions rather than the mere presence or absence of social contact are important for these outcomes in the current circumstances.^3^

In support of this, better quality social support acted as a moderator of the impact of adversity experiences on self-harm behaviours, which echoes research from before the current pandemic.^18,19^ Further, higher levels of loneliness exacerbated the impact of adversity experiences on self-harm behaviours. It is notable that the associations were specifically with self-harm *behaviours*; neither social support nor loneliness buffered the relationship between adversity experiences and *thoughts* about self-harm. Loneliness, low social support and adversities such as unemployment and financial problems are known risk factors self-harm and also for suicide, and the findings presented here confirm predictions from early in the COVID-19 pandemic that they would combine and exacerbate one another.^1,36^ Although in the current study this attenuation was modest, these results suggest the importance of available trusted others to provide understanding and support during pandemics, especially for individuals experiencing stressful life events.

Unexpectedly, however, this interaction was in the opposite direction for adversity *worries* and self-harm thoughts and behaviours. It is possible that people with higher levels of social support and lower loneliness talked about their worries more with others, with such conversations possibly leading to their worries being more prominent in their minds when they were then asked to self-report on them. This theory is supported by the fact that face-to-face contact exacerbated the relationship between adversity worries and self-harm thoughts, and telephone/video contact also exacerbated the relationship between adversity worries and self-harm *behaviours*.

Nonetheless, telephone/video calls still buffered the relationship between both adversity worries and experiences and self-harm *thoughts*. Future research could seek to disentangle whether the nature of telephone/video contact affects this moderation effect: it remains unclear whether these calls were made to friends/family, work colleagues, or telephone helplines such as the Samaritans. Prior findings using data from the same study as in the current analyses found that in the first month of the pandemic, small proportions of people who had reported self-harm thoughts (2.1%) or self-harm behaviours (4.6%) had utilised a helpline for mental health support, whilst around one-third had spoken with a friend or family member about their mental health.^12^

Analyses which disaggregated the social support measure into emotional and instrumental forms of support indicated that the quality of one’s emotional support, such as experiencing a lot of understanding and help from others and someone to talk to when feeling down, may be more important for self-harm thoughts than having someone to borrow something from or spend time with (instrumental support). However, there was evidence that instrumental support (not emotional support) buffered the association between experiencing adversities and self-harm behaviours. These findings are congruent with diathesis stress models of suicide risk which underscore the importance of having trusted others to rely on in the presence of near-term strain, worry, and adversity^6,7^ and suggest that public health campaigns that promote increase practical forms of support may help reduce suicide risk.^3^ These findings highlight the need for relatives, friends and neighbours to be encouraged to reach out to others who may be experiencing COVID-19 hardships such as unemployment, accessing essential items such as food or medicine, or contracting the virus itself.^3,36^

This study has a number of strengths as well as limitations. Strengths of this study include a long follow-up period with repeated measurements of predictor and outcome variables and the use of a large, well-stratified sample on demographic groups. Though data were weighted on the basis of population estimates of core demographics, sampling was not random and the findings can therefore not be generalised to the UK population as a whole. However, our goal was to identify associations between predictors and outcomes, and not to present population prevalence estimates. We also used a statistical modelling approach which accounted for time-invariant risk factors for self-harm such as genetic predisposition and adversity early in life^6,7^ and is thus an improvement upon prior research which did not account for these factors. This study also has several limitations. First, there were some differences in the wording of our measures of adversities and worries about adversities (see Table S3), and although selected to be congruent with one another, they may not therefore have captured the exact same adversity and worry.

Second, our measures of face-to-face/telephone/video contact lacked detail on who the participant was speaking with, and prior work from our research group suggests significant variability in the types of contacts accessed by people who report self-harm thoughts and behaviours.^12^ Third, our measure of self-harm behaviours did not specify what self-harming was, and participants may therefore have not reported behaviours they considered to be self-harming, but which may the behaviours clinically considered to fall under this umbrella label are diverse (e.g., self-poisoning or intentional destruction of bodily tissue). Fourth, we analysed data across 15 months, which included three different lockdowns when social support was largely provided virtually. It therefore remains unclear whether there were differences in the associations between the social factors we examined and outcomes depending on the precise social restrictions in place.

Our results demonstrate the importance during a pandemic of loneliness and social support for individuals, especially those who are facing adversities, highlighting their associations with self-harm thoughts and behaviours. Though modest, our moderation findings suggest that social support and loneliness help to buffer and exacerbate, respectively, against adversities. The provision of social support could therefore help to reduce the impact of pandemic-related adversities on self-harm. Whilst this study does not focus on suicide rates, self-harming is a strong risk factor for suicide risk, so helping to reduce risk factors for self-harming is an important mitigation strategy.^2,36^ It is therefore critical that policy makers and public health leaders not only focus on reducing adversities such as employment and financial hardship during the COVID-19 pandemic and potential future pandemics, but also develop community schemes that help to reduce loneliness and increase social support as part of self-harm and suicide prevention strategies. This is particularly important even as the pandemic abates as the detrimental impact of pandemic on self-harm and suicide is likely to accumulate and may even peak after the actual pandemic is under control.^4^

## Data Availability

The study protocol and user guide (which includes full details on recruitment, retention, data cleaning, and sample demographics) are available at https://github.com/UCL-BSH/CSSUserGuide.

## Acknowledgements

The researchers are grateful for the support of a number of organisations with their recruitment efforts including: the UKRI Mental Health Networks, Find Out Now, UCL BioResource, HealthWise Wales, SEO Works, FieldworkHub, and Optimal Workshop, as well as for Dr Liam Wright’s work on an earlier draft of this manuscript.

## Author Contribution

EP and DF conceptualised and designed the study. DF acquired funding, led the investigation, provided oversight on the methodology, provided software, and supervised the project. Data were curated, validated, and formally analysed by EP. EP wrote the original manuscript draft with input from DF, who then reviewed and edited the manuscript. Both authors approved the final version of the manuscript and had full access to and verified the data.

## Data Availability

Free-text data will not be made available due to stipulations set out by the ethics committee. The code used to run the analysis is available at https://osf.io/hmn9s/.

## Supplemental Materials

### Statistical Analysis

The basic fixed effects regression model can be expressed as follows:

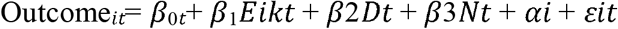

where Outcome_*it*_ is a measure of individual ‘s self-harm thoughts or self-harm behaviours at time *t*,E is individual *i*’s predictor variable at time *t*, D*t* is a vector of indicator variables for day, *Nt* is a continuous variable for days since lockdown, is unobserved time invariant confounding factors, and *ε* is error.

**Table S1:**
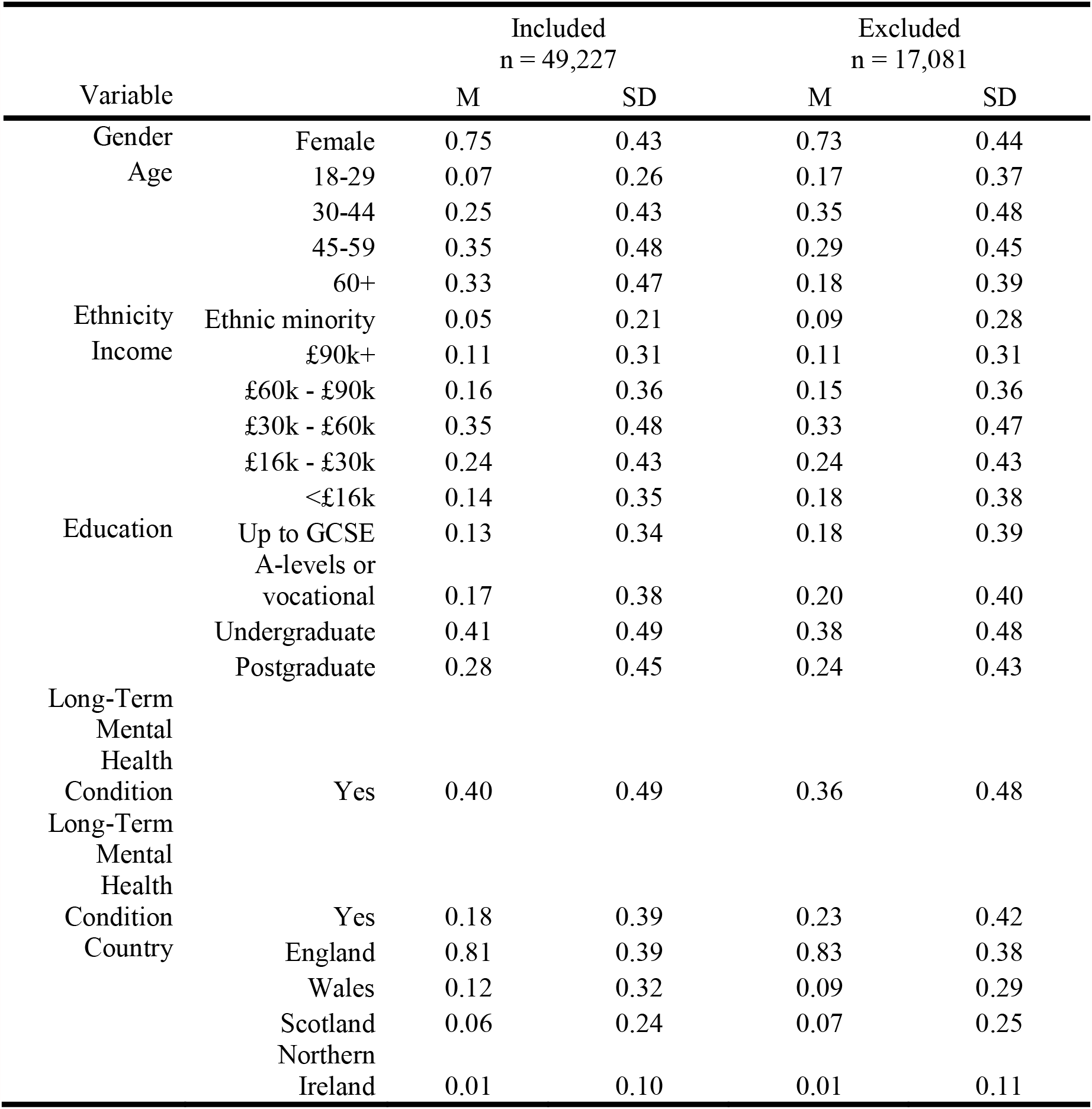
Characteristics of included and excluded participants, unweighted

## Measures

### Social Support

#### Perceived Social Support

Social support in the past week was measured using an adapted version of the six-item short form of Perceived Social Support Questionnaire (F-SozU K-6) ^29,30^. Each item is rated on a 5-point scale from “not true at all” to “very true”, with higher scores indicating higher levels of perceived social support. Minor adaptations made to question phrasing to make it relevant to experiences during COVID-19 can be found in Table S1.

**Table S2:** Adapted Perceived Social Support Questionnaire (F-SozU K-6)

| Original | Study adapted |
| --- | --- |
| I experience a lot of understanding and security from others | I have experienced a lot of understanding and support from others (emotional support) |
| I know a very close person whose help I can always count on | I have a very close person whose help I can always count on (emotional support) |
| If necessary, I can easily borrow something I might need from neighbours or friends | If necessary, I can easily borrow something I need from neighbours or friends (instrumental support) |
| I know several people with whom I like to do things | I have people with whom I can spend time and do things together (instrumental support) |
| When I am sick, I can without hesitation ask friends and family to take care of important matters for me | If I get sick, I have friends and family who will take care of me (instrumental support) |
| If I am down, I know to whom I can go to without hesitation | If I am feeling down, I have people I can talk to without hesitation (emotional support) |

#### Adversity experiences and adversity worries

Five categories of adversities were considered: finances, COVID-19 illness, social relationships, personal safety, and accessing essentials. We used multiple questionnaire items to measure each category (Table S2). Categories were treated as binary (absent vs. present) and were summed to create an index of the number of adversities experiences or worries at each time point. Adversity worries were captured from two multiple choice questions asked each wave: “Over the past week, have any of the following been worrying you at all, even if only in a minor way?”; “Have any of these things been causing you SIGNIFICANT stress? (e.g., they have been constantly on your mind or have been keeping you awake at night)”. The same response categories were given for each question.

**Table S3:** Questionnaire items used to measure adversity experiences and worries.

| Type of adversity | Adversity experiences | Adversity worries |
| --- | --- | --- |
| Financial | Lost your job/been unable to do paid work<br>Your spouse/partner lost their job or was unable to do paid work<br>Unable to pay bills/ rent/ mortgage<br>Major cut in household income (e.g., due to you or your partner being furloughed/ put on leave/ not receiving sufficient work)<br>Evicted/lost accommodation | Work (even if you feel your job is safe)<br>Losing your job / unemployment<br>Finances |
| COVID-19 illness | Currently have or previously had suspected or diagnosed COVID-19 | Catching COVID-19<br>Becoming seriously ill from COVID-19 |
| Social and relationship concerns | Somebody close to you is ill in hospital (due to COVID-19 or another illness)<br>You lost somebody close to you (due to COVID-19 or another cause) | Marriage or other romantic relationship<br>Friends or family living in your household<br>Friends or family living outside your household<br>Neighbours<br>Your pet |
| Accessing essentials | Unable to access sufficient food<br>Unable to access required medication | Getting medication<br>Getting food |
| Personal safety | Being physically harmed or hurt by somebody else<br>Being bullied, controlled, intimidated, or psychologically hurt by someone else | Your own safety / security |

### Variables used to describe the sample

All demographic and socio-economic variables were measured at baseline interview: country of residence (England, Scotland, Wales, Northern Ireland), gender (male, female), ethnicity (White, ethnic minority groups), age (18-29, 30-44, 45-59, 60+), annual income (< £16k, £16k - £30k, £30k - £60k, £60k - £90k, £90k +), and education level (up to GCSE, A-levels or equivalent, undergraduate degree, postgraduate degree).

Long-term physical health condition (yes, no) was assessed using a multiple-choice question on medical conditions. Included conditions were high blood pressure, diabetes, heart disease, lung disease, cancer, any other clinically-diagnosed chronic physical health conditions, or any disability. Long-term mental health condition (yes, no) was assessed with the same multiple choice question using items on clinically diagnosed depression, clinically diagnosed anxiety, and any other clinically diagnosed mental health problem.

**Table S4:** Descriptive statistics for entire sample and for those with variation in each outcome, weighted

|  |  | Total sample<br>n = 49,227 |  | Sample with variation in<br>self-harm thoughts<br>n = 11,559 |  | Sample with variation<br>in self-harm<br>behaviours<br>n = 3,740 |  |
| --- | --- | --- | --- | --- | --- | --- | --- |
| Variable |  | M | SD | M | SD | M | SD |
| Gender | Female | 0.52 | 0.50 | 0.55 | 0.50 | 0.55 | 0.50 |
| Age | 18-29 | 0.13 | 0.33 | 0.18 | 0.38 | 0.23 | 0.42 |
|  | 30-44 | 0.21 | 0.41 | 0.25 | 0.43 | 0.25 | 0.43 |
|  | 45-59 | 0.29 | 0.45 | 0.29 | 0.46 | 0.30 | 0.46 |
|  | 60+ | 0.38 | 0.49 | 0.28 | 0.45 | 0.21 | 0.41 |
| Ethnicity | Ethnic minority groups | 0.09 | 0.29 | 0.12 | 0.33 | 0.13 | 0.34 |
| Income | £90k+ | 0.08 | 0.27 | 0.06 | 0.24 | 0.05 | 0.22 |
|  | £60k - £90k | 0.12 | 0.33 | 0.11 | 0.31 | 0.08 | 0.27 |
|  | £30k - £60k | 0.33 | 0.47 | 0.30 | 0.46 | 0.26 | 0.44 |
|  | £16k - £30k | 0.28 | 0.45 | 0.28 | 0.45 | 0.26 | 0.44 |
|  | <£16k | 0.20 | 0.40 | 0.24 | 0.43 | 0.35 | 0.48 |
| Education | Up to GCSE | 0.31 | 0.46 | 0.30 | 0.46 | 0.33 | 0.47 |
|  | A-levels or vocational | 0.33 | 0.47 | 0.35 | 0.48 | 0.36 | 0.48 |
|  | Undergraduate | 0.22 | 0.41 | 0.21 | 0.40 | 0.18 | 0.39 |
|  | Postgraduate | 0.15 | 0.35 | 0.15 | 0.36 | 0.12 | 0.33 |
| Long-Term Mental<br>Health Condition | Yes | 0.44 | 0.50 | 0.48 | 0.50 | 0.53 | 0.50 |
| Long-Term Mental<br>Health Condition | Yes | 0.19 | 0.39 | 0.34 | 0.48 | 0.52 | 0.50 |
| Country | England | 0.84 | 0.36 | 0.85 | 0.36 | 0.84 | 0.36 |
|  | Wales | 0.06 | 0.23 | 0.05 | 0.22 | 0.05 | 0.23 |
|  | Scotland | 0.08 | 0.27 | 0.09 | 0.28 | 0.09 | 0.28 |
|  | Northern Ireland | 0.02 | 0.14 | 0.02 | 0.12 | 0.02 | 0.13 |

Note. Data were weighted to the proportions of gender, age, ethnicity, country, and education obtained from the Office for National Statistics.

**Table S5:** Sensitivity analysis: associations between disaggregated social support with self-harm thoughts and behaviours (main effects) derived from fixed effects logistic regression models.

| Variable | Self-Harm Thoughts |  |  | Self-Harm Behaviours |  |  |
| --- | --- | --- | --- | --- | --- | --- |
|  | OR | 95% CI |  | OR | 95% CI |  |
| Emotional Support | 0.53* | 0.51 | 0.55 | 0.79* | 0.74 | 0.83 |
| Instrumental Support | 0.76* | 0.74 | 0.79 | 0.77* | 0.72 | 0.81 |

*Note.* \* indicates $p < .05$ . Data were weighted to the proportions of gender, age, ethnicity, country, and education obtained from the Office for National

**Table S6:** Sensitivity analyses: associations between adversity and worries with self-harm thoughts and behaviours (main effects and interaction terms with the social support variable disaggregated) derived from fixed effects logistic regression models.

| Variable | Self-Harm Thoughts |  |  | Self-Harm Behaviours |  |  |
| --- | --- | --- | --- | --- | --- | --- |
|  | OR | 95% CI |  | OR | 95% CI |  |
| Adversity Experiences | 1.15* | 1.13 | 1.17 | 1.12* | 1.08 | 1.15 |
| Emotional Support | 0.47* | 0.46 | 0.48 | 0.69* | 0.66 | 0.73 |
| Interaction w/Emotional Support | 1.02* | 1.00 | 1.03 | 0.99 | 0.97 | 1.01 |
| Adversity Experiences | 1.12* | 1.10 | 1.14 | 1.08 | 1.05 | 1.12 |
| Instrumental Support | 0.55* | 0.53 | 0.56 | 0.68* | 0.65 | 0.72 |
| Interaction w/Instrumental Support | 0.99 | 0.98 | 1.00 | 0.96* | 0.94 | 0.98 |
| Adversity Worries | 1.43* | 1.39 | 1.46 | 1.33* | 1.27 | 1.38 |
| Emotional Support | 0.47* | 0.46 | 0.48 | 0.68* | 0.65 | 0.71 |
| Interaction w/Emotional Support | 1.02* | 1.00 | 1.04 | 1.04* | 1.01 | 1.07 |
| Adversity Worries | 1.40* | 1.37 | 1.43 | 1.30* | 1.24 | 1.35 |
| Instrumental Support | 0.55* | 0.54 | 0.57 | 0.67* | 0.64 | 0.71 |
| Interaction w/Instrumental Support | 1.01 | 0.99 | 1.03 | 1.02 | 0.99 | 1.05 |

*Note. \* indicates $p < .05$ . Data were weighted to the proportions of gender, age, ethnicity, country, and education obtained from the Office for National Statistics.*

